# Optimised oxygenation improves functional capacity during daily activities in patients with COPD on long-term oxygen therapy – a randomised crossover trial

**DOI:** 10.1101/2024.05.02.24306747

**Authors:** Linette Marie Kofod, Ejvind Frausing Hansen, Barbara Cristina Brocki, Morten Tange Kristensen, Nassim Roberts, Elisabeth Westerdahl

## Abstract

**Background:** Minimising hypoxemia during submaximal walking tests has a positive effect on exercise capacity and dyspnea in patients with COPD on long-term oxygen therapy (LTOT). However, the impact of optimising oxygenation during everyday tasks remains unexplored. Therefore, we investigated the effects of maintaining a target saturation on activities of daily living (ADL) using automated oxygen titration compared to conventional fixed oxygen flow.

**Methods:** In a double-blinded, randomised crossover trial, 31 patients with COPD on LTOT performed two GlittreADL tests to assess the functional capacity of everyday activities using a) their fixed oxygen dose and b) an adjusted flow from 0-8 L/min targeting a SpO_2_ of 90-94%. A closed-loop device automatically titrated the oxygen based on information from a Bluetooth wrist pulse oximeter.

**Results:** The patients reduced the time to perform the ADL-test by median (IQR) 38 (12–73) seconds, p<0.001, using automated titration compared to the fixed oxygen flow. The oxygen flow in the automated arm more than tripled to 5.4 (4.1–6.8) vs. 1.6 (1.1–2.1) L/min (fixed) during the test, p<0.001, while the time spent within SpO_2_-target was increased from 19% to 49%, p=0.002. Correspondingly, the patients experienced less dyspnea (BorgCR10); 5 (3–7) vs. 6 (4–8), p<0.001, in favour of the automated oxygen titration.

**Conclusions:** Improving oxygenation and extending the time spent within target saturation reduced dyspnea and improved functional capacity in activities of daily living in patients with COPD on LTOT.

**Trial registration number:** NCT05553847

## INTRODUCTION

Two landmark studies conducted more than 40 years ago, established the benefit of long-term oxygen therapy (LTOT) in terms of increased survival in patients with chronic obstructive pulmonary disease (COPD) who suffers from hypoxemic chronic respiratory failure.^1,2^ The aim of oxygen therapy is to provide patients with enough oxygen to bring the partial pressure of oxygen (PaO_2_) above 8 kPa at rest.^3^ However, during physical activity and exercises, the oxygen need increases, and the patients often experience periods of hypoxemia despite the use of LTOT.^3^ In a previous study by our group, the participants with COPD on LTOT spent 65% of the time during a walking test with a peripheral oxygen saturation (SpO2) <85%, when receiving their prescribed fixed oxygen dose.^4^

The consequences of intermittent hypoxemia seen in patients on LTOT on both patient-reported outcomes, functional capacity and mortality are poorly understood.^3,5^ Nevertheless, it is known that patients on LTOT have higher mortality, are very limited physically, and are less physical active than patients without the need of LTOT.^6^ Moreover, they have difficulties participating in activities of daily living (ADL) without desaturation and hypoxemia.^7^ Hypoxemia triggers dyspnea, which stands as the most debilitating symptom in the everyday life of patients with COPD,^8,9^ and the ability to engage in everyday tasks is essential to maintain an independent lifestyle.^9^ Yet, the influence of optimal oxygenation on ADL has not been investigated.

Closed-loop systems for oxygen delivery adjust the oxygen flow automatically based on input of oxygen saturation from a pulse oximeter attached on the patient’s finger. These closed-loop devices have shown to increase the time spent within a target saturation both during hospital admissions and in walking tests.^4,10-17^ The increased time within a target saturation during walking tests translates into meaningful improvements for patients with COPD on LTOT in terms of higher exercise capacity and alleviated dyspnea during walking.^4,17^ The movements during walking are uniform and repetitive, whereas during activities of daily living, the patient engages in diverse movements and may pause for rest. Daily activities such as dressing, showering, and cooking involve the use of both arms and legs, which amplifies ventilatory demands and dyspnea compared to walking only.^9^ This variability in the activities requires different levels of oxygen consumption, which could lead to significant fluctuations in the saturation.^7^ Closed-loop devices may be able to maintain a target saturation despite variability in oxygen needs. An aimable saturation is recommended to be above 90%, however, various guidelines seldom specify an upper saturation limit.^18^ The typical target for patients with stable COPD hovers around 90-94%.^2,4,17,19,20^ Targeting a specific saturation during everyday tasks is relevant due to the physiological effect of preventing hypoxemia, which could alleviate dyspnea during activities.

We hypothesised that by using a closed-loop system with automated oxygen flow fluctuating according to the actual oxygen demand we could optimise oxygenation, thereby enabling patients to efficiently accomplish daily tasks with reduced dyspnea.

The purpose of this study was to examine the effect of targeting a SpO_2_ of 90-94% on activities of daily living compared to the response of the usual fixed oxygen flow in a standardised ADL-test in patients with COPD on LTOT.

## METHODS

### Trial design and setting

This two-centre, double blinded, randomised crossover study enrolled patients with COPD and chronic respiratory failure with resting hypoxemia (PaO_2_ ≤ 7.3 kPa). Participants were recruited from the Departments of Pulmonology at Copenhagen University Hospital, Hvidovre and Copenhagen University Hospital, Bispebjerg-Frederiksberg, Denmark in the period from November 2022 to November 2023.

### Patients and recruitment

Inclusion criteria involved patients with clinically stable COPD who received LTOT according to the international criteria for home oxygen treatment,^3^ who were able to walk independently with or without walking aid and cognitively able to participate. Exclusion criteria were exacerbation in COPD treated with either antibiotics or prednisolone within the preceding three weeks or comorbidities known to impact physical functioning.

According to clinical routine patients on LTOT received a home care visit from a nurse specialised in oxygen treatment. In connection to the nurse visit, the patients’ medical records were screened, and eligible patients were invited to participate.

The included patients provided written informed consent before participation, and the study was approved by the Regional Research Ethics Committee (H-22032988) and the Danish Data Protection Agency j.nr. P-2022-625. The study was registered at ClinicalTrials.gov (NCT05553847), and the reporting follows the CONSORT statement for randomised crossover trials.

### Procedure

The patient’s ability to perform ADL was assessed using the GlittreADL test developed by Skumlien et al.^21^ On a single test day at the hospital, the patients conducted two ADL tests using A) their usual fixed oxygen dose and B) an automatically and continuously adjusted oxygen flow ranging from 0 to 8 L/min, set to achieve SpO_2_ in the target of 90-94%.

#### Activities of Daily living Assessment

GlittreADL is characterised as a semi-laboratory test validated in patients with COPD.^22,23^ It measures functional capacity (the maximum ability to perform activities) and reflects functional performance in daily physical activities known to be challenging for patients with COPD.^22,24^ A cut-off point of 3½ minutes in the GlittreADL test, has been proven effective in identifying patients with abnormal functional capacity, in terms of worse dyspnea, poorer health status, and decreased quality of life.^25^

Prior to the first test, baseline data was collected and the patients performed a practice round for familiarization.^23^ In the GlittreADL test, the patients rose from a seated position and walked 10 metre to a bookshelf, where they moved three 1 kg cartons from the top shelf at shoulder height to the middle shelf at hip height, down to the floor, and then stepwise back up to the top shelf. After turning, they walked back, sat down, and directly began the next lap. This was repeated for five laps. Rest was allowed during testing, but activity had to be resumed as soon as possible. Women and men carried a 2.5 and 5.0 kg backpack, respectively, throughout the test.^21^ To standardise the test all patients used a rollator (which also carried the oxygen cylinder and the closed-loop device), and the test was conducted without the usual two steps between the chair and the bookshelf (due to the use of rollator). Immediately after the test, the patients were asked to rate their level of dyspnea using Borg Dyspnea Scale CR10.^26^

#### Oxygen equipment

The oxygen supply came from the hospital’s 3-liter oxygen cylinder containing compressed oxygen. The cylinder was attached to the closed-loop device, O2matic HOT (O2matic ApS, Herlev, Denmark) and the equipment was placed on the rollator (figure 1). The patients wore a Nonin Wrist Pulse Oximeter (Nonin Medical, Inc., USA) which sent information on heart rate and oxygen saturation trough Bluetooth to the closed-loop device. In both arms the closed-loop device delivered the oxygen, and collected data every second on oxygen flow, SpO_2_ and heartrate. Data was transmitted to a cloud-based platform accessible only to the investigators. All patients wore the Optiflow™ (Fisher & Paykel) high flow nasal cannula for oxygen supplementation.

**Figure 1.**
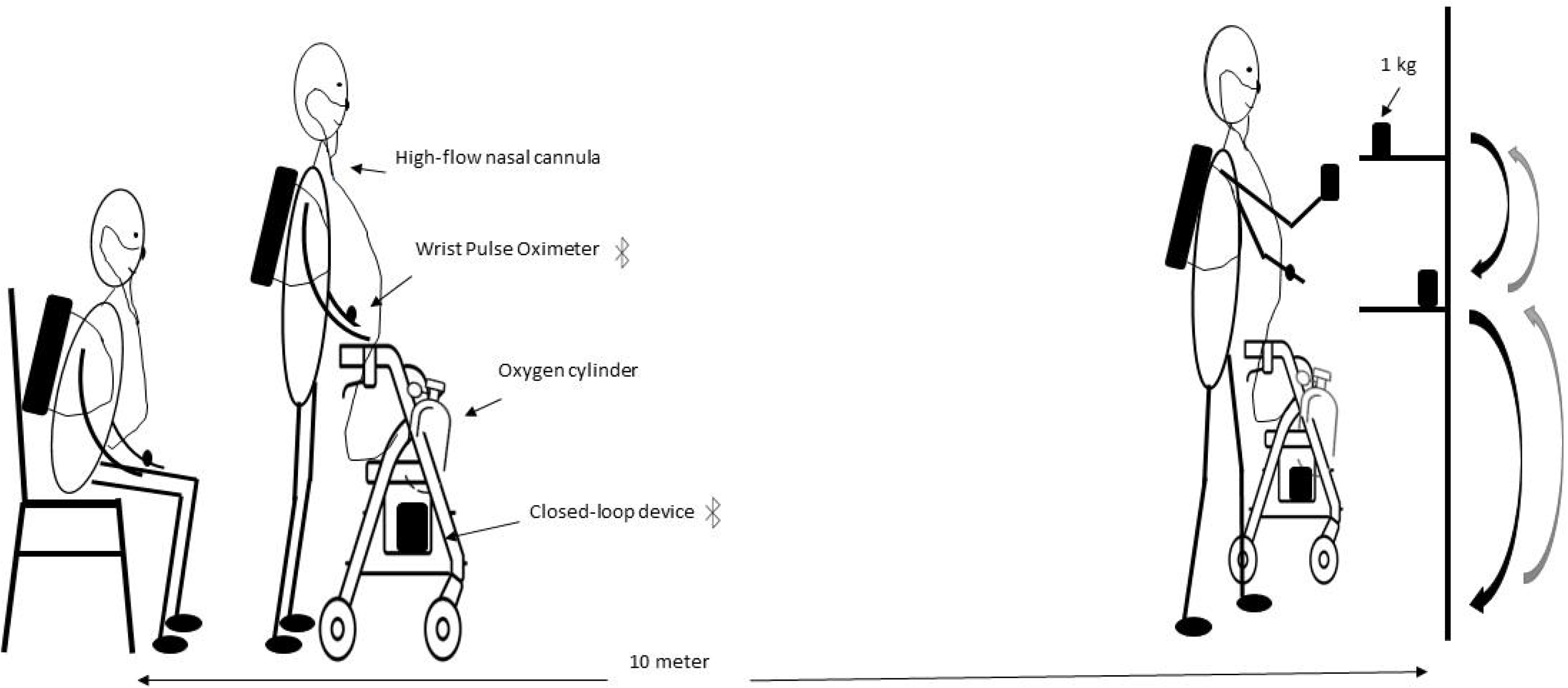
The GlittreADL test setup is identical in both arms. The patients carry a backpack, a pulse oximeter on the wrist, and uses a rollator. A high-flow nasal cannula is connected to the closed-loop device, which is attached to an oxygen cylinder. The patients rise from a chair, walk 10 m to a shelf, they move three 1 kg cartons, turn, walk back, and sit down. This is repeated five times in each arm.

*In the control arm*, the flow was kept fixed according to each patient’s medical prescription, if it was sufficient to maintain a SpO_2_>90% at rest.

*In the active arm*, the closed-loop device was set to aim at keeping the patients within a SpO_2_ target range of 90-94%. The oxygen flow was automatically adjusted up to 8 L/min based on the SpO_2_. The adjustments were done every second based on average SpO_2_ for the last 15 seconds.

### Outcomes

The primary outcome was difference between arms in time taken to complete the GlittreADL test. Secondary outcomes included the difference in Borg dyspnea score immediately after ending the GlittreADL test. Average oxygen flow during tests, differences in time spent within acceptable SpO_2_-interval (SpO_2_ 90–94%), time spent with moderate hypoxemia (SpO_2_ 85-89%) and with severe hypoxemia (SpO_2_<85%) were also assessed.

Demographics including body mass index, smoking status, spirometry, and comorbidities were collected, as well as the Medical Research Council dyspnea scale (MRC, range 1-5) and the COPD assessment test (CAT, range of 0-40).^27,28^

### Randomization and blinding

The patients were randomised after the familiarization of the test to either AB or BA. In AB arm, the patients received the usual fixed oxygen during the GlittreADL test first, followed by the target saturation titration, and vice versa in the BA arm. The crossover design was chosen to evaluate the response of the individual patient to different oxygen saturations. The randomization list was computer-generated compiled for each patient in REDCap electronic data capture tools (REDCap Consortium, Nashville, US) hosted at Capital Region of Denmark.

An independent person randomised and prepared the oxygen setup. The coding of the device to either fixed flow or automated titration was done in the cloud solution. The closed-loop device and the pulse oximeter was covered with black opaque tape to prevent visual observation. Both the assessor conducting the tests as well as the patient were blinded to the intervention. The minimum interval between test arms was 20 minutes to avoid carryover effect.^3^

### Statistical analysis

The sample size was determined based on the primary outcome, time to complete the GlittreADL test. The minimal important difference (MID) was 23 seconds and standard deviation expected to be 0.74 seconds.^25^ Based on alpha of 0.05 and a power of 80%, a power analysis revealed that a sample of 32 patients was needed to detect a statistical difference between arms.

MID in BorgCR10 dyspnea scale is one point difference in score.^29^

Continuous variables were examined for normality and analysed with either paired t-test (in case of normality) or Wilcoxon-signed-rank test (in case of non-normality). Pearson test was used for correlation. Test for carryover effect was performed by comparing the first and the second test using Wilcoxon test. IBM SPSS Statistics for Windows, ver. 28.0 was used for all statistical analyses. GraphPad Prism version 10.1.2 for Windows was used for figures.

## RESULTS

In the study period, 145 patients on LTOT were listed for nurse visits at Hvidovre Hospital. Out of these, 79 patients had a diagnosis of COPD and severe resting hypoxemia. After screening of medical records, 39 patients were eligible for the study, and 20 of them accepted participation. Additionally, patients were referred and included from Hvidovre Hospitals Pulmonary Rehabilitation Unit, and from Bispebjerg-Frederiksberg Hospital. Subsequently, 32 patients were included, as illustrated in figure 2. One patient was unable to complete the GlittreADL test due to severe dyspnea attack, significant desaturation, and the need of medical assistance, and was therefore excluded after randomisation. The baseline characteristics of the remaining 31 included patients are presented in table 1.

**Table 1.**
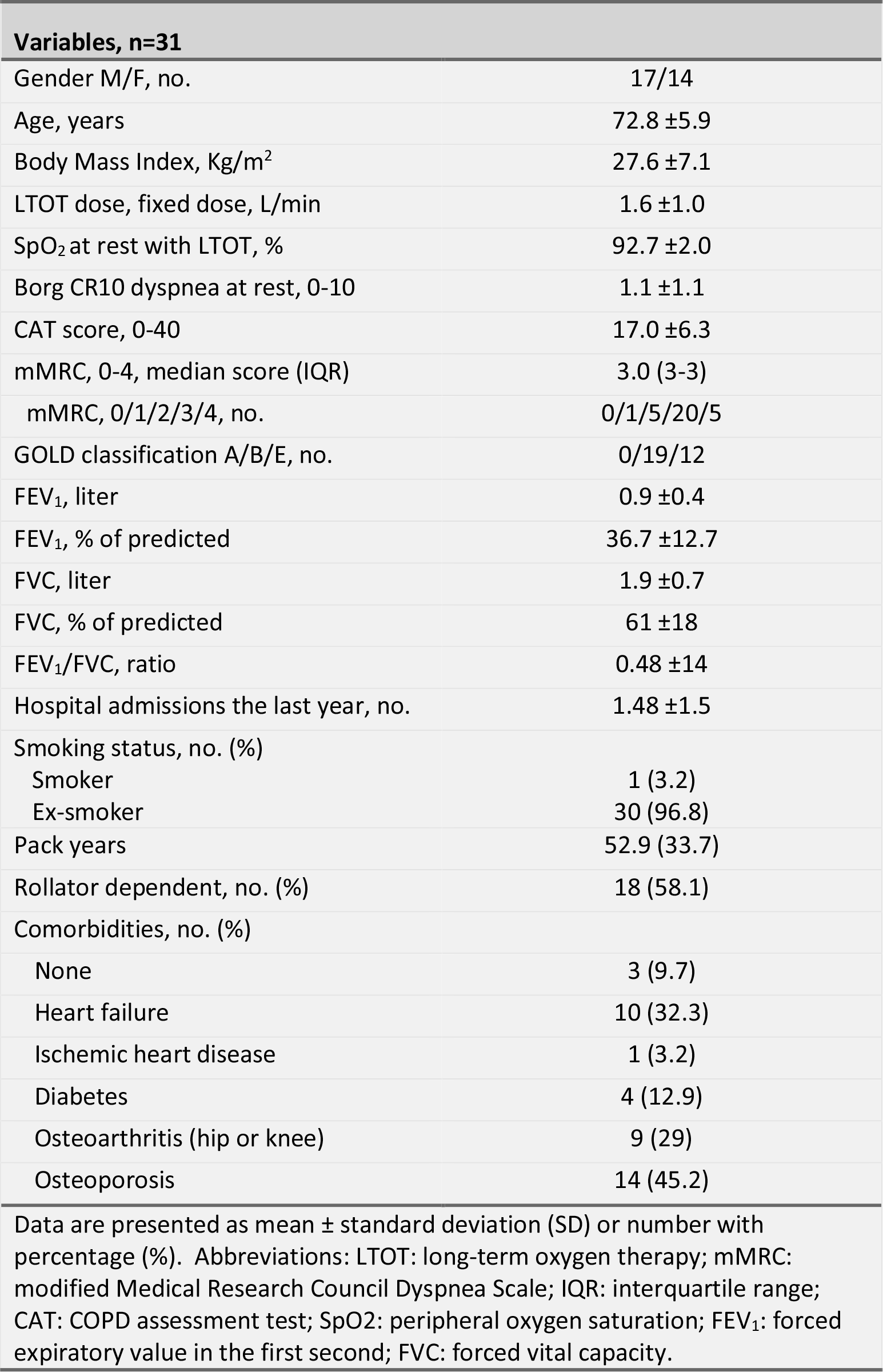
Characteristics of the study patients with COPD on LTOT.

| **Variables, n=31** |  |
| --- | --- |
| Gender M/F, no. | 17/14 |
| Age, years | 72.8 ±5.9 |
| Body Mass Index, Kg/m<sup>2</sup> | 27.6 ±7.1 |
| LTOT dose, fixed dose, L/min | 1.6 ±1.0 |
| SpO<sub>2</sub> at rest with LTOT, % | 92.7 ±2.0 |
| Borg CR10 dyspnea at rest, 0-10 | 1.1 ±1.1 |
| CAT score, 0-40 | 17.0 ±6.3 |
| mMRC, 0-4, median score (IQR) | 3.0 (3-3) |
| mMRC, 0/1/2/3/4, no. | 0/1/5/20/5 |
| GOLD classification A/B/E, no. | 0/19/12 |
| FEV<sub>1</sub>, liter | 0.9 ±0.4 |
| FEV<sub>1</sub>, % of predicted | 36.7 ±12.7 |
| FVC, liter | 1.9 ±0.7 |
| FVC, % of predicted | 61 ±18 |
| FEV<sub>1</sub>/FVC, ratio | 0.48 ±14 |
| Hospital admissions the last year, no. | 1.48 ±1.5 |
| Smoking status, no. (%) |  |
| Smoker | 1 (3.2) |
| Ex-smoker | 30 (96.8) |
| Pack years | 52.9 (33.7) |
| Rollator dependent, no. (%) | 18 (58.1) |
| Comorbidities, no. (%) |  |
| None | 3 (9.7) |
| Heart failure | 10 (32.3) |
| Ischemic heart disease | 1 (3.2) |
| Diabetes | 4 (12.9) |
| Osteoarthritis (hip or knee) | 9 (29) |
| Osteoporosis | 14 (45.2) |
| Data are presented as mean ± standard deviation (SD) or number with percentage (%). Abbreviations: LTOT: long-term oxygen therapy; mMRC: modified Medical Research Council Dyspnea Scale; IQR: interquartile range; CAT: COPD assessment test; SpO<sub>2</sub>: peripheral oxygen saturation; FEV<sub>1</sub>: forced expiratory value in the first second; FVC: forced vital capacity. |  |

**Figure 2.**
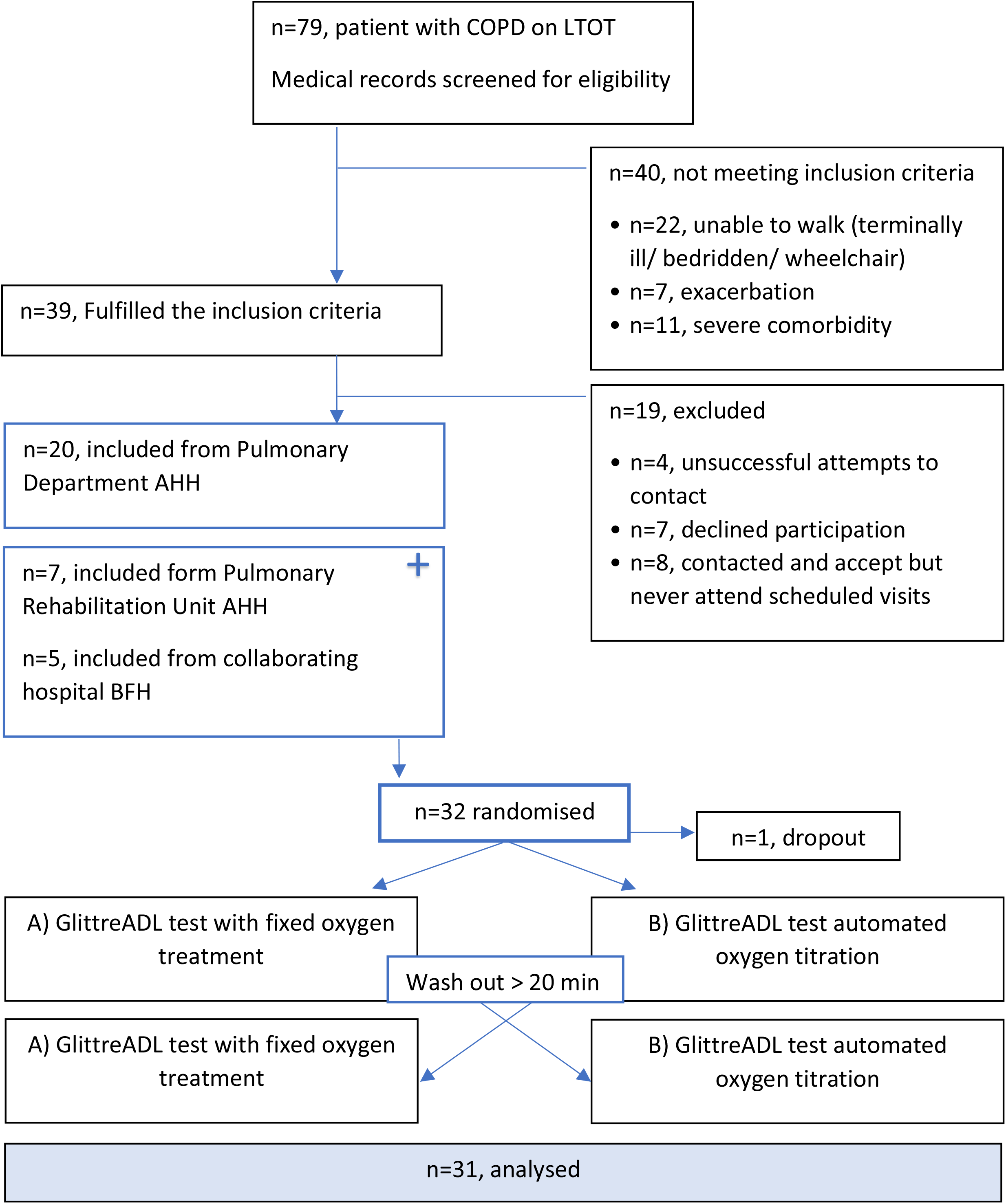
Flow diagram. LTOT: long-term oxygen therapy, AHH: Copenhagen University Hospital, Hvidovre; BFH: Copenhagen University Hospital, Bispebjerg-Frederiksbjerg.

Nine (29%) patients were unable to wear the backpack during the GlittreADL due to ankylosing spondylitis, severe osteoarthritis, clavicle/shoulder fracture or very severe dyspnea.

The primary outcome, time to complete the GlittreADL, was significantly reduced by median (interquartile range) 38 (12-73) sec, p<0.001, when using automated oxygen compared to the fixed dose, table 2. The corresponding difference in Borg dyspnea score was 1 (0-2), p<0.001 in favour of automated oxygen titration.

**Table 2.** Differences in outcomes between arms.

| **Table 2** Differences in outcomes between arms |  |  |  |  |
| --- | --- | --- | --- | --- |
| **Performance in the GlittreADL test, n=31** |  |  |  |  |
| **Variables** | **Fixed oxygen dose** | **Automated oxygen titration** | **Difference** | **P-value** |
| Time to complete GlittreADL, seconds | 427 (365–569) | 409 (300–502) | 38 (12–73) | <0.001 |
| Dyspnea, Borg CR10, score from 0-10 | 6 (4–8) | 5 (3–7) | 1 (0-2) | <0.001 |
| Time within target SpO<sub>2</sub> during ADL test, % | 18.7 (10.8–29.9) | 49.2 (20.3–72.4) | 21.4 (-3.2–52.6) | <0.001 |
| Median SpO<sub>2</sub> during test % | 86.6 (84.5–88.4) | 91.0 (86.9–92.0) | 3.4 (1.4–5.6) | <0.001 |
| Oxygen flow, L/min | 1.6 ±1.0 | 5.2 ±1.9 | 3.4 ±1.7 | <0.001 |
| Heart rate, bpm | 97.6 ±14.9 | 93.7 ±13.5 | 2.1 ±8.3 | 0.06 |
| Primary and secondary outcome measures during the GlittreADL test, when using usual fixed oxygen dose versus automated oxygen titration based on the peripheral oxygen saturation (SpO<sub>2</sub>). Target SpO<sub>2</sub>: 90–94%. Data presented as median with interquartile range or mean ±SD. Borg Dyspnea Scale CR10 at the end of the test. SpO<sub>2</sub>, oxygen flow and heart rate are presented as mean values throughout the test (n=28 in these 3 values). ADL: activities of daily living; L/min: litre per minutes; bpm: beats per minute. |  |  |  |  |

Twenty-seven (87%) patients exceeded the MID of 23 seconds and 1 point score, respectively, in either time to complete the GlittreADL or the Borg dyspnea score. Among these, 15 patients (48%) demonstrated improvements above the MID in both outcome measures. After excluding two outliers with improvements of 406 and 233 seconds, respectively, the time difference between GlittreADL tests remained at median 38 (9-63) seconds, p<0.001 (n=29). The fastest patient completed the GlittreADL in 3 minutes and 25 seconds using automated oxygen titration.

A significant larger proportion of time spent within target saturation was seen in the automated oxygen arm, p=0.002, figure 3. In more than 33% of the time in the ADL test, the patients experienced severe hypoxemia with an average SpO2<85%, when using the fixed oxygen dose. This time was significantly reduced to 17%, when adjusting the oxygen flow automatically, p=0.007. In the analysis testing for carryover effect, no statistical difference was found between test 1 and test 2, p=0.3.

**Figure 3.**
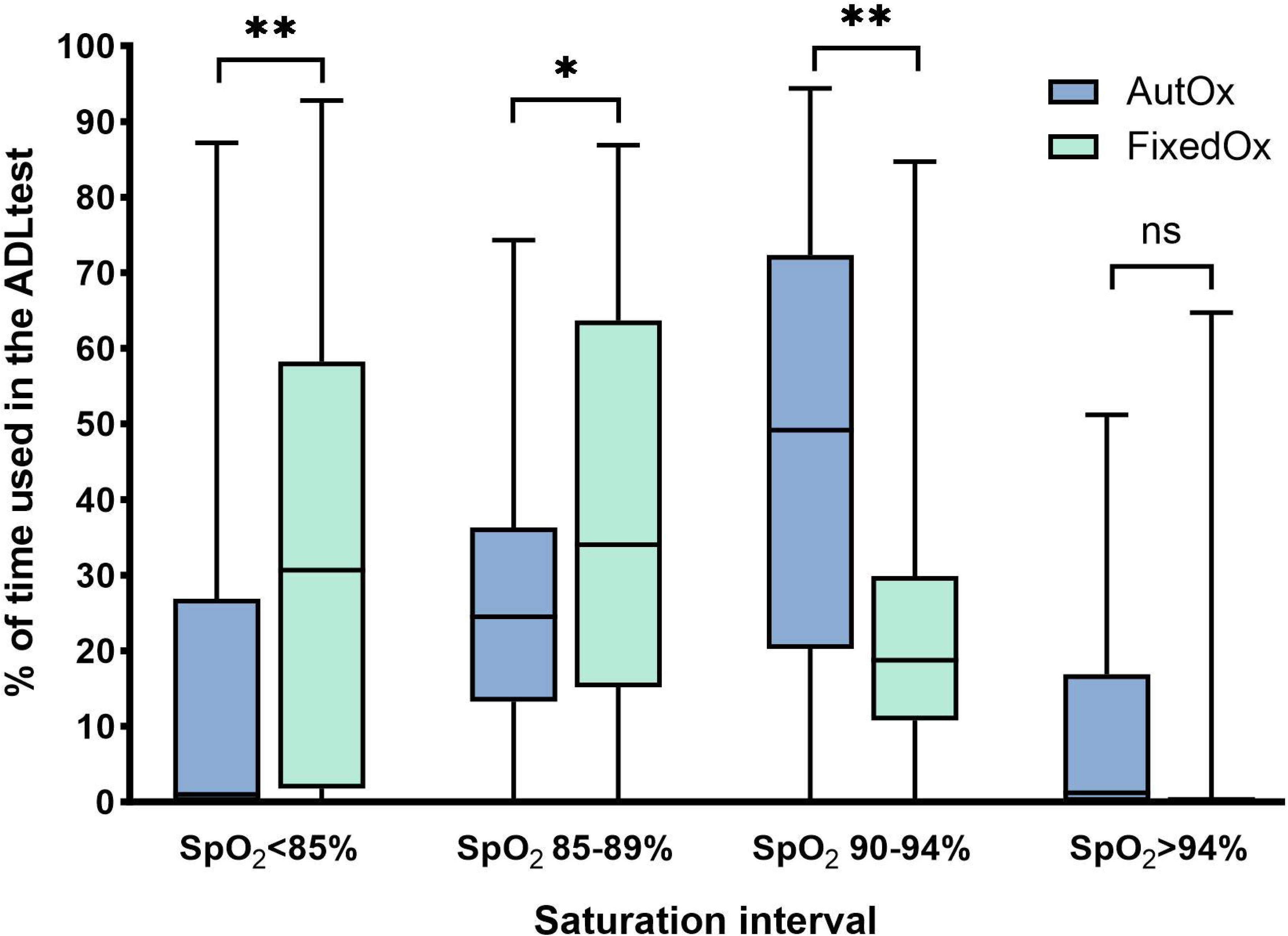
Percentage of time spend within saturation intervals. Boxplot showing median, interquartile range, minimum and maximum time spent in the different oxygen saturation (SpO2) intervals. X-axis: Four predefined oxygen saturations intervals. Y-axis: Percentage of the time taken to complete the GlittreADL test with Automated Oxygen titration (AutOx) and Fixed Oxygen dose (FixedOx). * p<0.05, ** p<0.01

A post hoc analysis excluding the two outliers, showed a correlation between degree of desaturation with fixed oxygen and the time difference between arms in completing the ADL test, r=0.43, p=0.017, figure 4.

**Figure 4.**
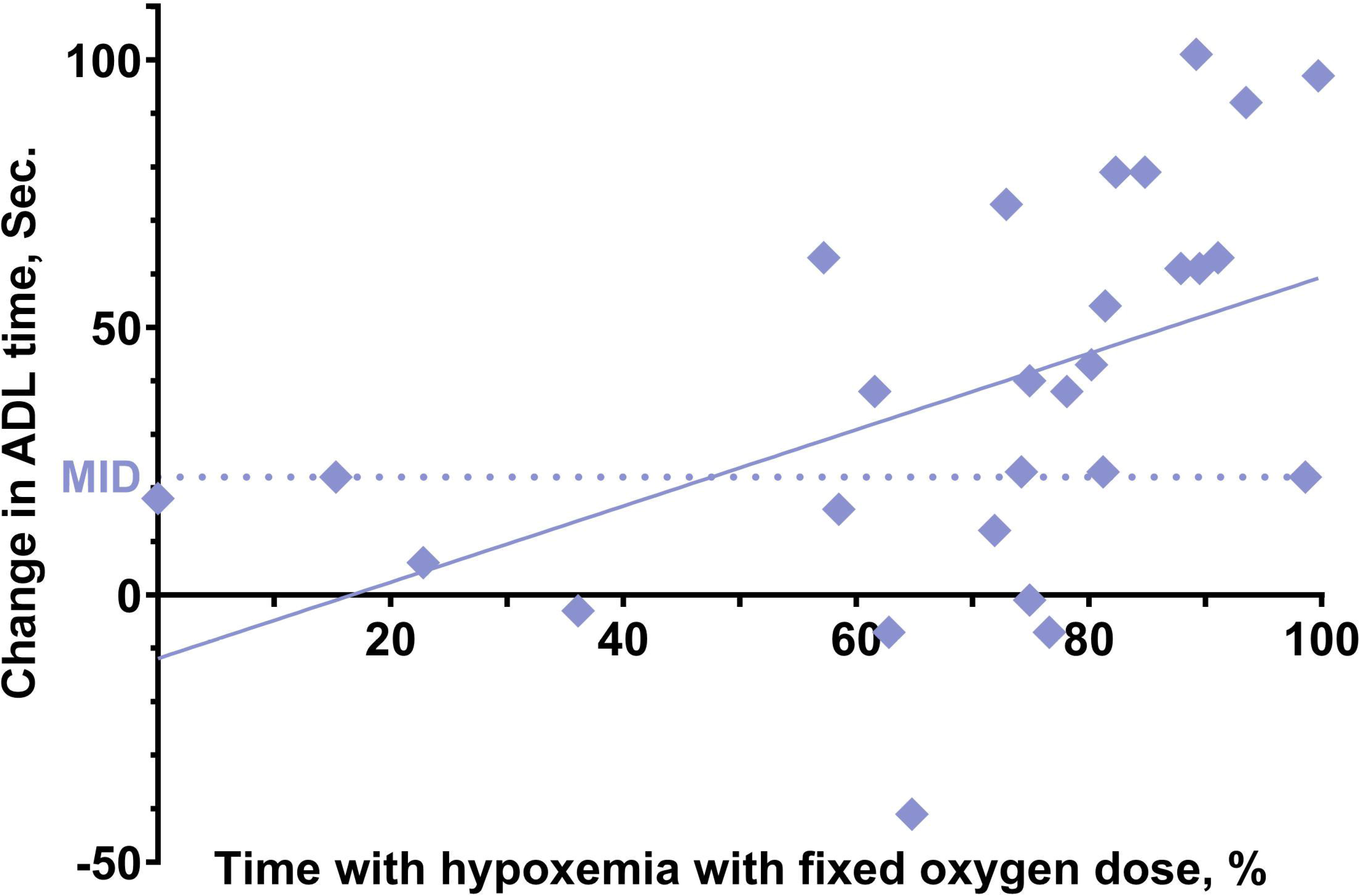
Time spent with hypoxemia relative to effect of optimised oxygenation. Each data point represents an individual patient, illustrating the percentage of time spent with hypoxemia (SpO_2_<90%) in the fixed oxygen arm relative to the change in performance of the GlittreADL test between arms. The minimal important difference (MID) in GlittreADL is 23 seconds illustrated with the dotted line.

## DISCUSSION

The present study showed that the capacity to perform activities of daily living improved in patients with COPD on LTOT when optimizing oxygenation. Correspondingly, the patients reported less dyspnea. The use of automated oxygen titration during the ADL-test increased time spent within a target saturation of 90-94% compared to the use of a fixed oxygen flow. However, it required more than three times as high an oxygen flow, which on average was 5.4 L/min in the adjusted flow arm compared to 1.6 L/min in the fixed flow arm.

To our knowledge, this is the first study to demonstrate the benefit of improved oxygenation on ADL performance. The physiological mechanism underlying this effect could involve the alleviation of dyspnea, as we also observed in the study, or it could be an improvement of skeletal muscle work through the minimisation of the peripheral desaturation. It is well known that patients with COPD are limited by dyspnea, peripheral muscle dysfunction and fatigue,^18^ and previous research has shown that supplementary oxygen reduce ventilatory demands,^30,31^ and improve muscle function.^32^ Patients requiring LTOT experience desaturation both during exercise and daily activities. Automated oxygen administration offers the opportunity to continuously adjust oxygen levels and thereby optimising the oxygen therapy not only by reducing periods with desaturation but stabilising the saturation as well.

The GlittreADL test involves various functional tasks, such as walking, sitting down, rising from a chair, crouching, and moving objects. In our study, the patients took an average of 7.3 minutes to complete the test. We removed the steps from the original test to ensure that all patients began under the same conditions, and in order to use a rollator. It can be speculated that patients would spend less time on the test without the need to climb stairs. However, only one patient managed to complete the test in less than the cut-off points of 3½ minutes, which is the discrimination between normal and abnormal functional capacity. This emphasizes that our study participants were severely limited in daily activities due to their condition. The observed improvement of 38 seconds, while statistically significant, also holds clinical relevance.^25^ This improvement could significantly impact patients’ daily lives by potentially facilitating activities such as showering, cooking, mobility, and social engagement.

The patients had the option to sit after each lab until ready to continue. Although patients were encouraged to remain seated for as short a time as possible between laps, this rest period probably allowed them to recover in terms of heart rate, dyspnea intensity, and also in terms of oxygen saturation. Compared to our previous study,^4^ the improvement in dyspnea and performance was less pronounced in the present study. This difference could be due to the nature of the two tests: the Endurance Shuttle Walk Test being a submaximal test with constant work, and the GlittreADL being more comparable to real-life situations with the possibility to pause and with a self-chosen pace. Additionally, the potential maximum oxygen flows were higher in the previous study, with 15 L/min compared to the maximum of 8 L/min in the present study. The reason for this difference was that we wanted to simulate the patients’ situation at home, where flows of 15 L/min is typically not possible. All in all, 87% of the participating patients, in the present study, had a clinically relevant improvement in either the time to complete the ADL task or on dyspnea. However, it required tree time as high an oxygen flow and still the patients were within SpO2-target for only 47% of the time.

We found a correlation between time spent with SpO_2_<90% during the test using fixed oxygen, and the improvement in time to complete the test, when oxygen was adjusted to achieve a target interval of 90-94% (figure 4). In other words, the more the patients desaturated the more they profited of the adjusted oxygen flow. This strengthens the hypothesis that it is the correction of hypoxemia that leads to an increase in ADL performance.

Currently, the primary rationale for providing patients on LTOT with portable oxygen devices for ambulatory oxygen is the possibility to prolong the time spent with LTOT.^3,5^ Lacasse et al. showed in 2005 that patients were not more physical active when receiving ambulatory oxygen versus no ambulatory oxygen.^33^ However, the saturation levels of the participating patients were not assessed. It could be argued that if the patients desaturated while moving outdoor, they were still very limited by hypoxemia, thus lacking capacity to move. Our study demonstrated an effect of oxygen therapy on ADL and could consequently support the argument that ambulatory oxygen is beneficial for patients, especially if it could be personalised based on the specific task ahead of them.

The actual impact on ADL needs to be examined in a home setting, which highlights a limitation of our study: the GlittreADL, being a simi-laboratory test, is only a surrogate for the challenges patients encounter in their home environment, and it may not fully capture all aspects of functional capacity relevant to everyday activities in patients with COPD. Using additional outcome measures or assessments could have provided a more comprehensive understanding of functional capacity. Our study involved a relatively small sample size of 31 patients, which coukld limit the generalisability of the findings.

We chose to modify the GlittreADL test to ensure inclusion of patients in need of a walking aid, which was 58% of the patients (table 1). We did not find any studies describing the use of the GlittreADL without steps, leading us to conclude that all studies have involved patients capable of walking without a rollator and climbing stairs without assistance. This modification of the otherwise validated test represents a clear limitation of our study. Nevertheless, since the patients served as their own controls, we maintain confidence in the results. Also, some patients were unable to carry the backpack during the test, mainly due to severe comorbidity. However, in this case, a study showed validity and comparable SpO_2_ responses between GlittreADL with and without backpack.^34^ Last, the closed-loop device and Bluetooth pulse oximeter may have technical limitations or inaccuracies that could affect the reliability of the measurements.

A strength of our study was the blinding of the patients with the closed-loop device used for both fixed flow and variable flow, and an Optiflow nasal cannula, typically utilised in an inpatient hospital setting. The Optiflow minimised the risk of the patients noticing an increased flow, and they did not report any burning sensation in the nose as experienced in our earlier study.^4^ Further, the crossover design with both tests conducted on the same day ensured that the patients were in similar health status and that the results very well reflect the individual response to the optimisation of the oxygen level.

In conclusion, the functional capacity in activities of daily living in patients with COPD on LTOT improved, when improving oxygenation and extending the time spent within target saturation. Concurrently, the patients reported of less dyspnea with enhanced oxygenation. Future studies should focus on the possibilities and benefits of improving oxygenation and maintaining a target saturation when moving outdoor and doing activities at home.

## Data Availability

All data produced in the present study are available upon reasonable request to the authors

## Acknowledgements

The authors acknowledge all the patients for their time and energy. Special thanks to Maria Swennergren Hansen og Kira Marie Skibdal for their assistance with randomisation, preparing equipment, and their helpfulness throughout the study.

## Contributors Study design

LMK, EFH, BCB, MTK and EW. Patient recruitment and data collection: LMK, EFH and NR. Analysing or interpreting data: LMK, EFH, MTK and EW. LMK prepared the manuscript. All authors have critically revised, read, and approved the manuscript. LMK takes the responsibility for the integrity.

## Data availability statement

Data is available upon reasonable request.

## Ethics approval

This study involves human participants and was approved by the Regional Research Ethics Committee of the Capital Region of Denmark (H-22032988) and the Danish Data Protection Agency j.nr. P-2022-625. Participants gave informed consent to participate in the study before taking part.

